# Reliability, validity and dimensionality of the GHQ-12 in South African populations: Structural equation modelling (SEM)

**DOI:** 10.1101/2023.06.06.23290967

**Authors:** Clement Nyuyki Kufe, Colleen Bernstein, Kerry Wilson

## Abstract

**Introduction:** Health Care Workers (HCWs) were among the high-risk groups for SARS-CoV-2 infection and suffer a high burden of poor mental health including depression, anxiety, traumatic stress, avoidance and burnout. The 12-Item General Health Questionnaire (GHQ-12) has showed best fit in both a one-factor structure and a multidimensional structure for the screening of common mental disorders and psychiatric well-being. The aim was to test for the reliability and validity and ascertain the factor structure of the GHQ-12 in a South African HCW population.

**Methods:** Data was collected from 832 public hospital and clinic staff during the COVID-19 pandemic in Gauteng, South Africa. The factor structure of the GHQ12 in this professional population was examined by exploratory factor analysis (EFA) to identify factors, confirmatory factor analysis (CFA) for construct validity and structural equation modelling (SEM).

**Results:** The GHQ-12 median score was higher (25) in women than in men (24), p=0.044. The determinant for the correlation matrix was=0.047, the Barlett test of sphericity was p<0.001, Chi square=2086.9 and Kaiser-Meyer-Olkin (KMO) of sampling adequacy was 0.86. The four factors identified were labelled as Social-Dysfunction (37.8%), Anxiety-Depression (35.4%) Capable (24.9%) and Self-Efficacy (22.7%). The entire sample had a Cronbach’s alpha of 0.85, with 0.69 for Social-Dysfunction, 0.74 for Anxiety-Depression, 0.64 for Capable and 0.52 for Self-Efficacy in orthogonal (varimax) factor loadings.

**Conclusions:** The GHQ-12 tool displayed adequate reliability and validity in measuring psychological distress in a professional group with a four-factor model suggesting multidimensionality in this group rather than a unidimensional construct.

## Introduction

The 2019 coronavirus disease (COVID-19) arose in China in December 2019 and by early February 2020 reports of psychological pressures on Chinese Health Care Workers (HCWs) were published (1). COVID-19 did not only affect the mental health of Chinese HCWs it also affected the physical health of HCWs as it rapidly spread throughout the world. By the 11^th^ of March the World Health Organization (WHO) had declared COVID-19 a global pandemic (2). The first recognised COVID-19 case was diagnosed in South Africa on the 5^th^ of March and 116 cases registered by the 18^th^ March. Following a National State of Disaster declared on the 15^th^ of March, with schools closing on the 18^th^ of March, a country-wide lockdown began on the 26^th^ March 2020 in South Africa. HCWs were designated essential services and continued working at the forefront in the fight against COVID-19 pandemic. HCWs were responsible for screening, testing and managing COVID-19 positive patients as well as unknown COVID-19 patients, along with those responsible for cleaning health care facilities and, dealing with the administration and management of the facility (3). A high degree (57.4%) of psychological distress and a strong association between perceived risks associated with the presence of COVID-19 in the health care workplace and psychological distress was reported in HCWs in South Africa during the pandemic (3).

Epidemic outbreaks have been known to place an unpreceded demand of HCWs resulting in increased deaths, infection, shortage of medication and vaccines, increase workload and lack of personal protective equipment, feelings of inadequate support all exacerbating the mental distress and burden of HCWs (3–5). A systematic review of the mental health of HCWs in the COVID-19 pandemic reported the lowest prevalence’s of depression, anxiety and stress of 24.1% and 29.8% in HCWs…(country) and highest prevalence of 55.9% (country), 67.6% (country) and 62.9% (country) for cross-sectional studies published in English (6). Another review of studies at the time identified risk factors for COVID-19 –related health impact in HCWs as: inadequate hand hygiene, high-risk department, diagnosed family member, suboptimal had hygiene before and after contact with patients, improper personal protective equipment use, close contact with patients (≥12 times/day), long daily contact hours (≥ 15 hours) and weakness with female HCWs disproportionately affected (2).

Due to stigma, HCWs were likely to suffer psychological distress in silence which may have led to an increased risk of suicidal ideation (7–9). The uneasy climate during COVID-19 characterised by panic and fear in the general population was likely increased in HCWs as they needed to manage patient care at a time when their own physical and mental wellbeing was at stake (10). The unremitting stress of a pandemic or epidemic could trigger psychological issues of anxiety, fear, panic attacks, posttraumatic stress symptoms, psychological distress, stigma and avoidance of contact, depressive tendencies, sleep disturbances, helplessness, interpersonal social isolation from family social support and concern regarding contagion exposure to their friends and family (11–13). At the same time, programmes put in place faced resistance from HCWs in acknowledging the experience of psychological difficulties (14). There is an urgent need to provide evidenced-informed data on the adverse mental health and well-being effects of HCWs to mitigate these challenges and provide recommendations for resilience (12)

The General Health Questionnaire (GHQ) developed in the 1970s was initially intended for use as a unidimensional tool of 60-items to describe the risk of mental health disorders. Many shortened versions of the GHQ have been developed such as the GHQ-30, the GHQ-28, the GHQ-20 and the GHQ-12 (15–18), all of which have been subject to factor analytic procedures to identify whether each of the scaled versions give additional utility (19). The shorter GHQ-12 has been used in epidemiologic studies, the majority of which and subsequent factor analytic studies have failed to accept the unitary construct (20) and proposed a multidimensional structure (21). Because of its brevity the 12-item GHQ has been widely used as a screening instrument for depression, mental illness, anxiety/depression and depression disorder in many countries (15,22–27). While the GHQ has been widely used to date, to the researchers’ knowledge no study has examined the factor structure on a South African sample. The factor analytic approach assesses interrelationships within a set of variables to construct a smaller number of hypothetical variables or factors that contain the essential information of the larger set of observed variables, thus reducing the overall complexity of the data by taking advantage of the inherent interdependencies (28). Factor analysis is useful for measuring constructs that are not readily observable, summarising large observations into smaller number and providing evidence of construct validity by hypothesis testing (19). Further, studies that have been carried out to assess the mental health of HCWs including the use of the 12-item General Health Questionnaire (GHQ-12), have not been validated in all professional populations. It would thus be important to validate, test the reliability of the GHQ-12 and describe the factor structure in South African populations, more specifically on a professional sample of HCW’s. This study aimed at assessing the validity, reliability and determine the factor structure of the GHQ-12 in the South African, Gauteng, health care worker population.

## Methods

### Study design and population

A cross sectional study was conducted in three tertiary hospitals, two regional hospitals, two community health centres and nine clinics in Gauteng, South Africa from August to October 2020. Data was collected from 832 male and female hospital staff during the COVID-19 pandemic using a self–administered questionnaire exploring socio-demographic information, assessing stress and psychological effects, perceptions, attitudes and behaviour around COVID-19. The GHQ-12 consisted of 12 statements to which respondents indicated agreement on a four-point scale (1= Better than usual, 2=Same as usual, 3=Worse than usual and 4=Much worse than usual) on mental stress (18). The detailed description of the study and instruments have been done elsewhere (3).

### Data management and analysis

Data was captured on Research Electronic Data Capture (REDCap) (29) and analysed using Stata version 16.1/MP (StataCorp, College Station, TX, USA) after exploration to establish missing data points. The univariate normality of the items was assessed using the skewness and kurtosis of the responses. A series of factor analyses (both exploratory and confirmatory factor analyses) were implemented. Factor analysis was used to examine the theory driven factor structure as evidential criteria using exploratory factor analysis (EFA) (30–32) and by testing the assumptions using confirmatory factor analysis (CFA) to indicate which model should be chosen and compared with (33–35). Using EFA, the appropriate factors to explain the relationship between observed variables was established. Orthogonal (varimax) was employed to determine the number of factors to retain by using the following criteria: 1) eigenvalues of the factors ≥ 1, 2) Cattell’s scree test, 3) internal consistency and 4) factors that yielded meaningful psychological constructs. Structural equation modelling (SEM) assessed the predictive utility of the factors identified with CFA by using the following goodness-of-fit criteria: Likelihood ratio, Population error, Information criteria, Baseline comparison and Size of residuals.

The study was approved by the University of Witwatersrand, Human Research Ethics Committee (Medical) Clearance certificate No: M2006103. Written consent was obtained from all participants before completing the anonymous online questionnaire.

## Results

The median (25^th^ – 75^th^ percentile) age of the respondents was 44 (34–54) years. Women formed the largest group of participants at 89.9% and 3.9% indicated other for gender. The majority, 99%, were South African citizens with the remainder from Southern Africa. The most common home language was isiZulu followed by Setswana.

The median GHQ score (1-4 coding) was 25 for women higher when compared to men who scored 24, p=0.044. The determinant for the correlation matrix was=0.034, the Barlett’s test of sphericity was p<0.001, Chi square=2262.171 and Kaiser-Meyer-Olkin (KMO) of sampling adequacy was KMO=0.877, the variables were inter-correlated enough to warrant factor analysis. The entire sample had a general average inter-item covariance of 0.33 and a Cronbach’s alpha of 0.85, 0.85 for men and 0.84 for women illustrating satisfactory internal consistency in both groups and in men and women respectively. The range of the item-scale correlation was 0.50 for playing a useful part in things to 0.69 for the unhappy and depressed in the entire sample (Table 1). The values of Cronbach’s alpha of 0.85 and the McDonald’s omega of 0.85 were comparative/comparable. The majority of respondents completed all twelve questions, although “Been able to enjoy normal day to day activities” was the most skipped question.

**Table 1:** Adjusted Item-Scale Correlation (AISC) and internal consistency of the 12-items of the GHQ questions.

| **GHQ-Items** | **n** | **AISC** | **$\alpha$** |
| --- | --- | --- | --- |
| 1. Been able to **concentrate** on what you are doing? | 847 | 0.59 | 0.84 |
| 2. Lost much **sleep** over **worry**? | 836 | 0.61 | 0.84 |
| 3. Felt that you are playing a **useful** part in things? | 830 | 0.50 | 0.85 |
| 4. Felt **capable** of making **decisions** about things? | 831 | 0.57 | 0.84 |
| 5. Felt constantly under **strain**? | 846 | 0.66 | 0.84 |
| 6. Felt that you could not overcome your **difficulties**? | 838 | 0.68 | 0.83 |
| 7. Been able to **enjoy** your normal day-to-day **activities**? | 829 | 0.61 | 0.84 |
| 8. Been able to **face** up to your **problems**? | 844 | 0.61 | 0.84 |
| 9. Been feeling unhappy and **depressed**? | 846 | 0.69 | 0.83 |
| 10. Been **losing confidence** in yourself? | 850 | 0.68 | 0.83 |
| 11. Been thinking of yourself as a **worthless** person? | 849 | 0.59 | 0.84 |
| 12. Been feeling reasonably **happy** all things considered? | 832 | 0.62 | 0.84 |

Item-scale analysis of the 12-Item Global Health Questionnaire
AISC: Adjusted Item-Scale Correlation. $\alpha$ examines reliability by determining the internal consistency of a test or average correlation of items (variables) within the test.

### Reliability, validity and the factor structure

For the factor analysis, 672 participants were included, 4 factors were retained, and all had a coefficient greater than 0.30. The test of internal consistency represented by a scale of reliability coefficient was 0.85 for all the 12 items, 0.78 for factor I, 0.76 for factor II, 0.64 for factor III and 0.64 for factor IV in orthogonal (varimax) rotation. The reliability and validity was adequate for use in this population. The factor loadings were labelled as follows: factor I labelled Social-Dysfunction (37.8%); factor II called Anxiety-and-Depression (35.4%); factor III called Capable (22.7%), factor IV labelled Self-Efficacy (24.9) with the first 3 factors accumulatively representing 98.1% (Table 2).

**Table 2:**
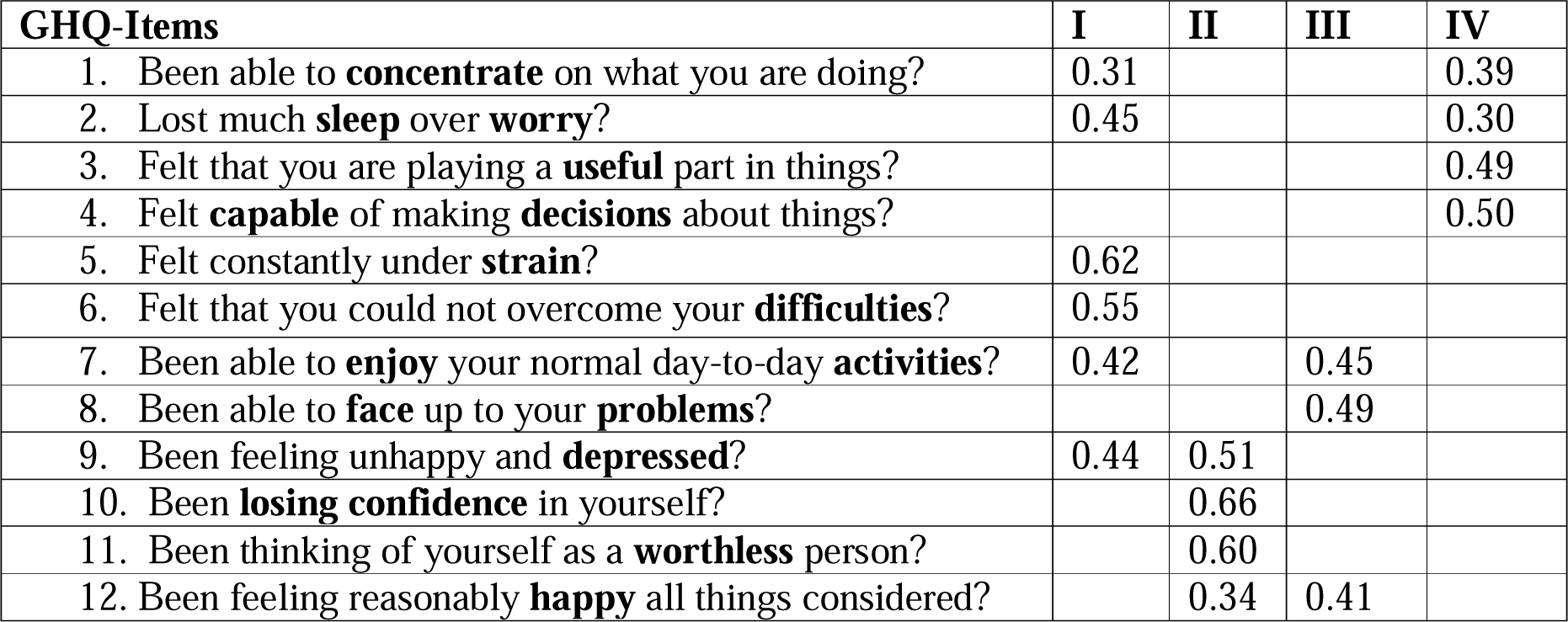
Maximum likelihood estimates for orthogonal (varimax) factor loadings of the GHQ-12

### Model fit, comparison and predictive utility

CFA was used to ascertain whether the factor structure that was developed from the data matched the chosen conceptual models and an analysis Goodness-of-fit test between the data and the model (36). Model fit was evaluated by examining the size and statistical significance of the factor loadings as well as comparative fit index (CFI), Tucker-Lewis index (TLI), the standardized root mean square residual (SRMR) and the root mean square error of approximation (RMSEA) at 90% confidence interval (CI). Accordingly, a RMSEA of 0.01, 0.05 and 0.08 is accepted for excellent, good and mediocre fit respectively at 90% CI (37) while a SRMR of less than 0.08 is generally considered a very good fit and 0.05–1.0 are considered acceptable fit (38). A CFI and TLI close to 0.95 represents a good fit between hypothesised model and the data while values in the range of 0.90–0.95 are acceptable (38–40). Competing models were compared by use of the Akaike’s information criterion (AIC), the Bayesian information criterion (BIC) and the likelihood-ratio test where the lower values for AIC and BIC indicate a better fit.

We examined the size and statistical significance of the factor loadings and common goodness-of-fit statistics. A unidimensional model was tested for all the 12 items in Model 1 named mental distress. In Model 2, a four-factor structure was tested and labelled as Social dysfunction (4 items), Anxiety and Depression (4 items), Self-Efficacy (2 items) and Capable (2 items). While in Model 3, the four-factor structure was maintained but item distribution was Social dysfunction (3 items), Anxiety and Depression (3 items), Self-Efficacy (3 items) and Capable (3 items). Structural equation modelling (SEM) assessed the predictive utility of the factors identified with CFA by using goodness-of-fit criteria (Table 3). The best fitting model was model 3 with the most acceptable values (Figure 1) and the highest overall coefficient of determination (CD) or R-squared as 0.972 (Supplementary files 1).

**Figure 1:**
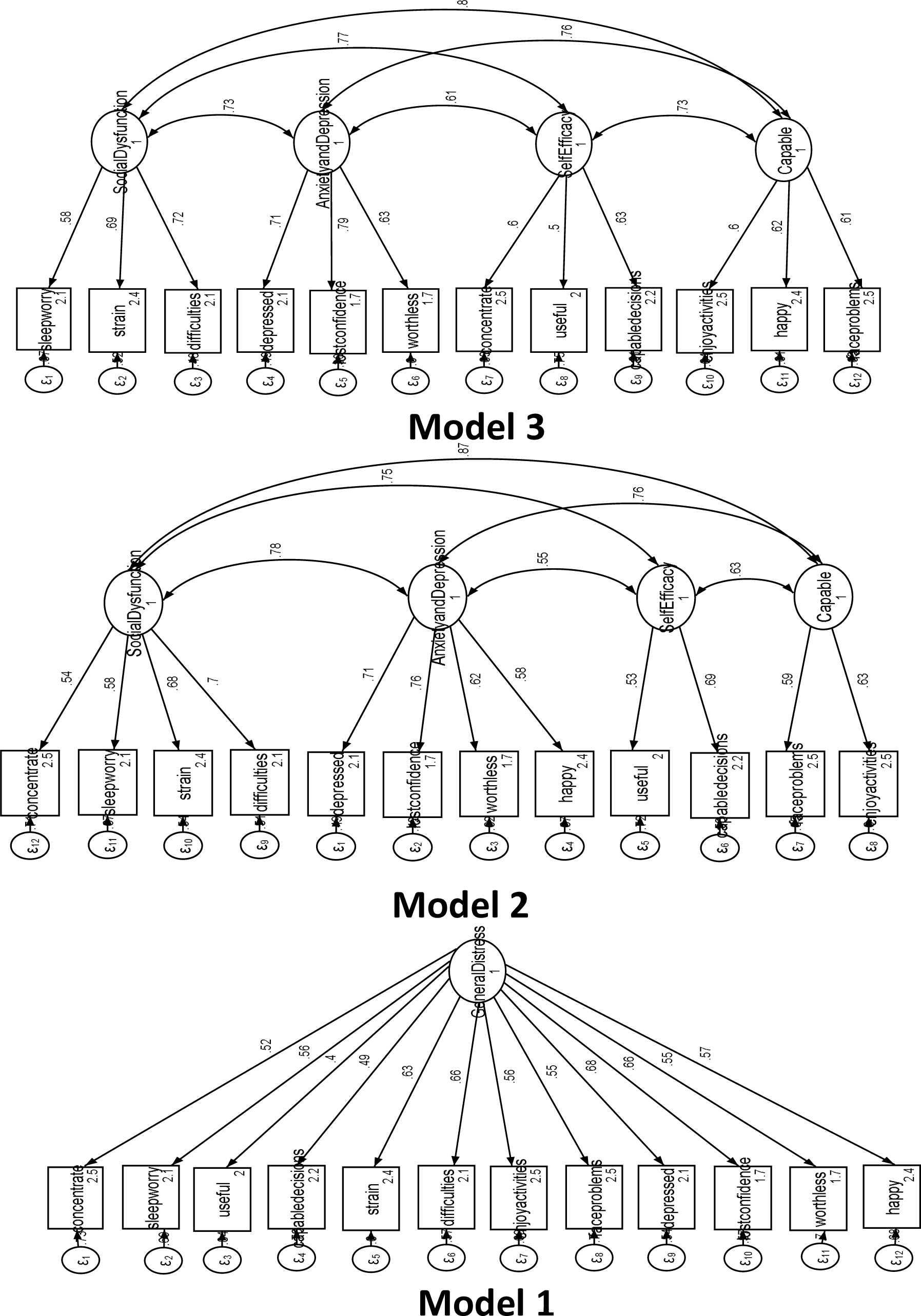
Factor models for the GHQ-12 of HCWs in South Africa

**Table 3:**
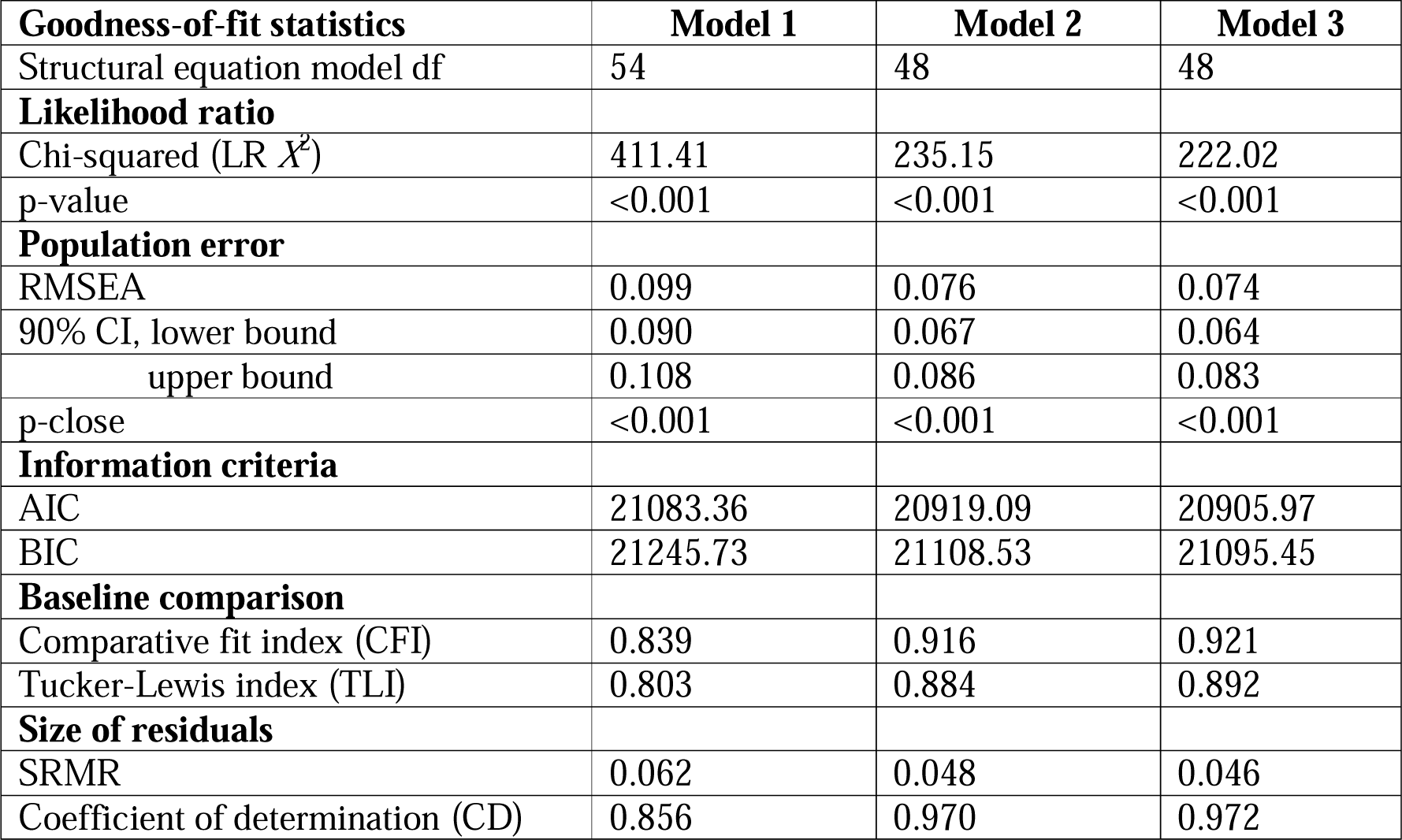
Goodness-of-fit statistics for tested models of the GHQ-12 in HCWs of South Africa LR *X*^2^: likelihood ratio chi-square, df: degrees of freedom, CFI: Comparative fit index, LTI: Tucker-Lewis index (TLI), SRMR: standardized root mean square residual, RMSEA: root mean square error of approximation, CI: confidence interval.

The retained results from the SEM are presented in Model 3 of Figure 1. Social dysfunction was positively and statistically significantly associated with lost much sleep over worry, felt constantly under strain, felt that you could not overcome your difficulties and been able to concentrate on what you are doing. Anxiety and Depression was positively and strongly associated with been feeling unhappy and depressed, been losing confidence in yourself and been thinking of yourself as a worthless person. Self-Efficacy was positively and statistically significantly associated with felt that you are playing a useful part in things and felt capable of making decisions about things. Capable was associated and statistically significantly with been able to enjoy your normal day-to-day activities and been able to face up to your problems.

## Discussion

To our knowledge, this is the first evaluation of the factor structure of the GHQ-12 in a South Africa population. This study in South African HCWs showed good reliability and validity of the GHQ-12 in a professional South African population. The factor structure was not unitary (one factor distress) but a multidimensional scale with a robust external validity for all four factors identified.

This study demonstrated adequate reliability and validity for a professional South African population and demonstrated the best-fit data with multi-dimensional structure similar to a multicentre study involving Ethiopian populations (41) and to the literate Kenyan study (42). The Cronbach’s alpha and the coefficient omega of McDonald are measures of the composite reliability for the GHQ-12 and demonstrated a reliability higher than 0.70. The McDonald’s coefficient omega is unbiased with congeneric items with uncorrelated errors unlike the Cronbach’s alpha (43,44) and the two estimators were comparative in this study. Similar findings of another study in a random sample of Brazilian physicians registered in the medical Council system (45) and a similar Cronbach’s alpha was found in primary care patients of Indonesians (46).

Unlike the unidimensional facture structure reported in an Indonesian population and as originally developed (46) and the bi-factor structure in the Brazilian study (45) as well as the Norwegian Navy (47), our study suggested a four-factor structure similar to the multidimensionality reported in the Spanish population (48). In the SEM we did not include insomnia and mental health problems (HSCL-25) as in unidimensional domains (47,49) but opted for the multidimensionality as obtained from the initial results of the EFA and CFA.

This study showed a four-factor model as the best explanation in a sample of South African HCWs as compared to the three-factor model labelled as Anxiety-Depression, Social Dysfunction and Loss of Confidence when testing for the six-factor analytic models (50) and reported in a longitudinal and cross-sectional studies (51–53). This study confirmed the valid use of GHQ-12 in a professional occupational group as previously showed in a study of the young civil servants in China but differs in the suggested factor structure which was three-factor as compared to the four-factor structure in our study (53). A Finnish study using GHQ-12 and GHQ-20 concluded that the GHQ-12 had a three-factor structure and the GHQ-20 had a four-factor structure which was superior to the GHQ-12 as it provided an additional factor named anhedonia suggesting some discriminative power (54). The above studies that suggested a three-factor structure provided little information beyond that of a general factor while our study showed more information with the four-factor structure after SEM. The three-factor structure was basically similar to our four-factor structure with similar loadings but with differences in items that load on each factor. The main difference was observed in the factor orderings such that, the 3 items in our factor II (Anxiety and Depression) were the same as 3 of the 4 items in factor II and 2 of the items in our factor I (Social Dysfunction) were similar to the 2 of 3 items in factor III (48).

Additionally, the data was explored for modification indices (MI) suggesting covariance between “Been losing confidence in yourself?” and “Been thinking of yourself as a worthless person?” (MI:40.87), “Been feeling unhappy and depressed?” and “Been thinking of yourself as a worthless person?” (MI:26.51), and “Felt constantly under strain?” and “Been thinking of yourself as a worthless person?” (MI:27.87) (Supplementary files 2). This may be explained by the existence of an unspecified factor not included in the model that might partially account for the relationship between the variables or measurement artefacts. The existence of between-factor differences suggest that the GHQ-12 has multidimensional characteristics not captured by a severity score as reported by Vanheule and Bogaerts in a Belgian sample and Graetz in an Australian sample of young people (55,56).

### Strengths and limitations

The GHQ-12 is a short tool and is easily scored, and has been previously used in different cultures for screening purposes to detect psychological distress. The validity of the GHQ-12 in South African HCWs was assessed by the Barlett’s test supplemented by the Kaiser-Meyer-Olkin (KMO) as a measure of sampling adequacy. The use of EFA permitted a compromise by balancing the parsimony and comprehensiveness in the model that contains just enough factors to explain the important variations in the measured variables and later using multiple methods to advise on the plausible and appropriate factor solutions (30,32,34,57–60). The EFA is a theory-testing model suggesting hypotheses, while the CFA based on strong theoretical foundation explicitly tests the hypotheses about the factor structure, and serves as construct validity. The interpretation of the factors is complicated because of lack of prior knowledge leading to difficulty in interpretation of the results obtained from EFA. However, the use of CFA and structural equation modelling with standardised coefficients permitted the confirmation of the factors. The longer versions of the GHQ are useful in assessing the degree of psychological morbidity and outcomes for clients managed at mental services. The primordial strength of this study is the validation of the utility of the GHQ-12 in South African HCWs. The limitation of this study include the non-generalisability of the results to the whole South African and African population for screening as this study was limited to HCWs only.

## Conclusions

The GHQ-12 displayed adequate reliability and validity in measuring psychological distress of HCWs. The factor structure suggested multidimensionality rather than a unidimensional construct. The findings of this study affirm the effectiveness of the GHQ-12 in a professional group of South Africans. The GHQ-12 can be a useful screening instrument in the South African population for general symptoms of mental distress to effectively assess overall psychological well-being and detect non-psychiatric problems. Further research is warranted to test the reliability and validity of the GHQ-12 in the general South African population.

## Data Availability

All data produced in the present study are available upon reasonable request to the authors

## Acknowledgements

We are grateful to the Health Care Workers (HCWs) who consented and participated in this study and the Gauteng Department of Health (GDOH) for the assistance.

## Contributions

CNK analysed the data, drafted and revised the manuscript. CB and KW conceived the study. CNK is responsible for the overall content as the guarantor. All authors read and approved the final version of the manuscript.

## Funding

The study received funding from the National Institute for Occupational Health (NIOH), Johannesburg, South Africa and the University of British Columbia Canada.

## Notes

### Competing Interest Statement

The authors have declared no competing interest.

### Funding Statement

The study received funding from the National Institute for Occupational Health (NIOH) and the Gauteng Department of Health (GDOH), Johannesburg, South Africa.

