## Supplementary files 1 for "Reliability, validity and dimensionality of the GHQ-12 in South African populations: Structural equation modelling (SEM)"

**Supplementary Files 2: Structural Equation Modelling**

SEM 1: Equation-level goodness of fit standardised

| **Dependent variable** | **fitted** | **Variance predicted** | **R-squared** |
| --- | --- | --- | --- |
| 1. Been able to concentrate on what you are doing? | 0.779 | 0.205 | 0.263 |
| 2. Lost much sleep over worry? | 1.050 | 0.319 | 0.304 |
| 3. Felt that you are playing a useful part in things? | 0.872 | 0.144 | 0.165 |
| 4. Felt capable of making decisions about things? | 0.892 | 0.193 | 0.217 |
| 5. Felt constantly under strain? | 1.137 | 0.468 | 0.411 |
| 6. Felt that you could not overcome your difficulties? | 1.053 | 0.470 | 0.447 |
| 7. Been able to enjoy your normal day-to-day activities? | 1.045 | 0.353 | 0.338 |
| 8. Been feeling unhappy and depressed? | 1.262 | 0.587 | 0.465 |
| 9. Been able to face up to your problems? | 0.786 | 0.216 | 0.275 |
| 10. Been losing confidence in yourself? | 1.165 | 0.456 | 0.391 |
| 11. Been thinking of yourself as a worthless person? | 1.179 | 0.312 | 0.265 |
| 12. Been feeling reasonably happy all things considered? | 0.919 | 0.267 | 0.290 |
| Overall |  |  | 0.856 |

SEM 2: Equation-level goodness of fit standardised

| **Dependent variable** | **fitted** | **Variance predicted** | **R-squared** |
| --- | --- | --- | --- |
| 1. Been able to concentrate on what you are doing? | 0.779 | 0.216 | 0.277 |
| 2. Lost much sleep over worry? | 1.050 | 0.343 | 0.326 |
| 3. Felt that you are playing a useful part in things? | 0.872 | 0.268 | 0.308 |
| 4. Felt capable of making decisions about things? | 0.892 | 0.424 | 0.475 |
| 5. Felt constantly under strain? | 1.137 | 0.542 | 0.477 |
| 6. Felt that you could not overcome your difficulties? | 1.053 | 0.541 | 0.514 |
| 7. Been able to enjoy your normal day-to-day activities? | 1.045 | 0.500 | 0.478 |
| 8. Been able to face up to your problems? | 0.786 | 0.268 | 0.341 |
| 9. Been feeling unhappy and depressed? | 1.261 | 0.689 | 0.546 |
| 10. Been losing confidence in yourself? | 1.165 | 0.642 | 0.551 |
| 11. Been thinking of yourself as a worthless person? | 1.179 | 0.426 | 0.361 |
| 12. Been feeling reasonably happy all things considered? | 0.920 | 0.275 | 0.299 |
| Overall |  |  | 0.970 |

SEM 3: Equation-level goodness of fit standardised

| **Dependent variable** | **fitted** | **Variance predicted** | **R-squared** |
| --- | --- | --- | --- |
| 1. Been able to concentrate on what you are doing? | 0.779 | 0.273 | 0.350 |
| 2. Lost much sleep over worry? | 1.050 | 0.339 | 0.322 |
| 3. Felt that you are playing a useful part in things? | 0.872 | 0.233 | 0.267 |
| 4. Felt capable of making decisions about things? | 0.892 | 0.325 | 0.365 |
| 5. Felt constantly under strain? | 1.137 | 0.567 | 0.499 |
| 6. Felt that you could not overcome your difficulties? | 1.053 | 0.576 | 0.547 |
| 7. Been able to enjoy your normal day-to-day activities? | 1.045 | 0.447 | 0.428 |
| 8. Been able to face up to your problems? | 0.786 | 0.284 | 0.362 |
| 9. Been feeling unhappy and depressed? | 1.262 | 0.678 | 0.537 |
| 10. Been losing confidence in yourself? | 1.165 | 0.690 | 0.592 |
| 11. Been thinking of yourself as a worthless person? | 1.179 | 0.441 | 0.374 |
| 12. Been feeling reasonably happy all things considered? | 0.920 | 0.318 | 0.346 |
| Overall |  |  | 0.972 |

**Supplementary Files 2: Modification indices**

|  | MI | P>MI | EPC | Standard EPC |
| --- | --- | --- | --- | --- |
| cov(e.lostconfidence,e.worthless) | 40.87 | 0.00 | 0.248 | 0.419 |
| cov(e.depressed,e.worthless) | 26.51 | 0.00 | -0.200 | -0.305 |
| cov(e.strain,e.worthless) | 27.87 | 0.00 | -0.159 | -0.246 |

MI: Modification Indices, EPC: Expected Parameter Change
